# Clustering of predicted loss-of-function variants in genes linked with monogenic disease can explain incomplete penetrance

**DOI:** 10.1101/2023.10.11.23296535

**Authors:** Robin N. Beaumont, Gareth Hawkes, Adam C. Gunning, Caroline F. Wright

## Abstract

Predicted loss-of-function variants (pLoFs) are often associated with disease. For genes linked with monogenic diseases, we hypothesised that pLoFs present in apparently unaffected individuals may cluster in LoF-tolerant regions. We compared the distribution of pLoFs in ClinVar versus 454,773 individuals in UK Biobank and clustered the variants using Gaussian mixture models. We found that genes in which haploinsufficiency causes developmental disorders with incomplete penetrance were less likely to have a uniform pLoF distribution than other genes (P<2.2x10-6). In some cases (e.g., *ARID1B* and *GATA6*), pLoF variants in the first quarter of the gene could be rescued by an alternative translation start site and should not be reported as pathogenic. In other cases (e.g., *ODC1*), pathogenic pLoFs were clustered only at the end of the gene, consistent with a gain-of-function disease mechanism. Our results support the use of localised constraint metrics when interpreting variants.

## Introduction

Contrary to expectation, many individuals in the population harbour predicted loss of function (pLoF) variants in genes where haploinsufficiency is known to cause highly penetrant monogenic conditions^1–3^. For example, pLoF variants in genes that cause severe developmental disorders (DD) in childhood would not be expected to be present at appreciable levels in the general adult population. Nonetheless, we and others have previously shown that thousands of individuals in UK Biobank (UKB) carry pLoF variants in DD genes and have phenotypes consistent with incomplete penetrance or reduced expressivity, though very few individuals have DD diagnoses^4–7^. There are several possible explanations for this observation. One possibility is that genetic or environmental modifiers alter the impact of individual variants^8^, such that the penetrance of pathogenic variants identified in affected families or disease cohorts may be over-estimated. An alternative explanation is that some pLoF variants in these genes do not cause loss of function, either because they are technical false positives^9^ or mosaic variants, or because they can be rescued through a variety of mechanisms, including alternative transcription^10^, exon skipping^11^, escape from nonsense-mediated decay (NMD)^12^, and translation re-initiation^13^. It is important to distinguish between benign pLoF variants that produce near-normal levels of functional protein, and pathogenic variants that result in substantially reduced protein product, both for estimating penetrance and interpreting diagnostic results.

Constraint metrics derived from population variation have been extremely useful for identifying genes that are intolerant to pLoF variation^14,15^, and regions of genes that are intolerant to missense variation^16,17^. However, the location of pLoF variants in genes has not been systematically investigated at large scale due to lack of sequence data on large numbers of individuals. We used cluster analysis of exome sequence (ES) data from UKB to identify genes showing distinct patterns in the location of pLoF variants. We then investigated genes showing these distinct profiles of pLoF location to determine whether they could explain the presence of such putatively pathogenic variants in a population cohort.

## Results

### pLoF variants are more likely to be non-uniformly distributed than missense or synonymous variants

We calculated the relative location of every coding variant detected in ES data from 454,773 individuals in UKB in the coding sequence (CDS) of each gene, and the relative proportion of variants grouped by consequence class in each transcript^18^ (synonymous, missense, pLoF) within each quintile of the CDS. We then used Gaussian mixtures to cluster the profile of the variants of each class within the transcripts into seven clusters (Fig. 1). Of these, three clusters represented variants being distributed more-or-less uniformly throughout the CDS (clusters 1-3; Fig. 2), and one identified genes with no variants of a particular class (not shown). The remaining three clusters showed distinct patterns in the location of variants, with at least one quintile of the CDS containing zero (or very few) variants of a given variant class, and most variants either being towards the first or second half of the gene (clusters 4-6; Fig 2).

**Figure 1:**
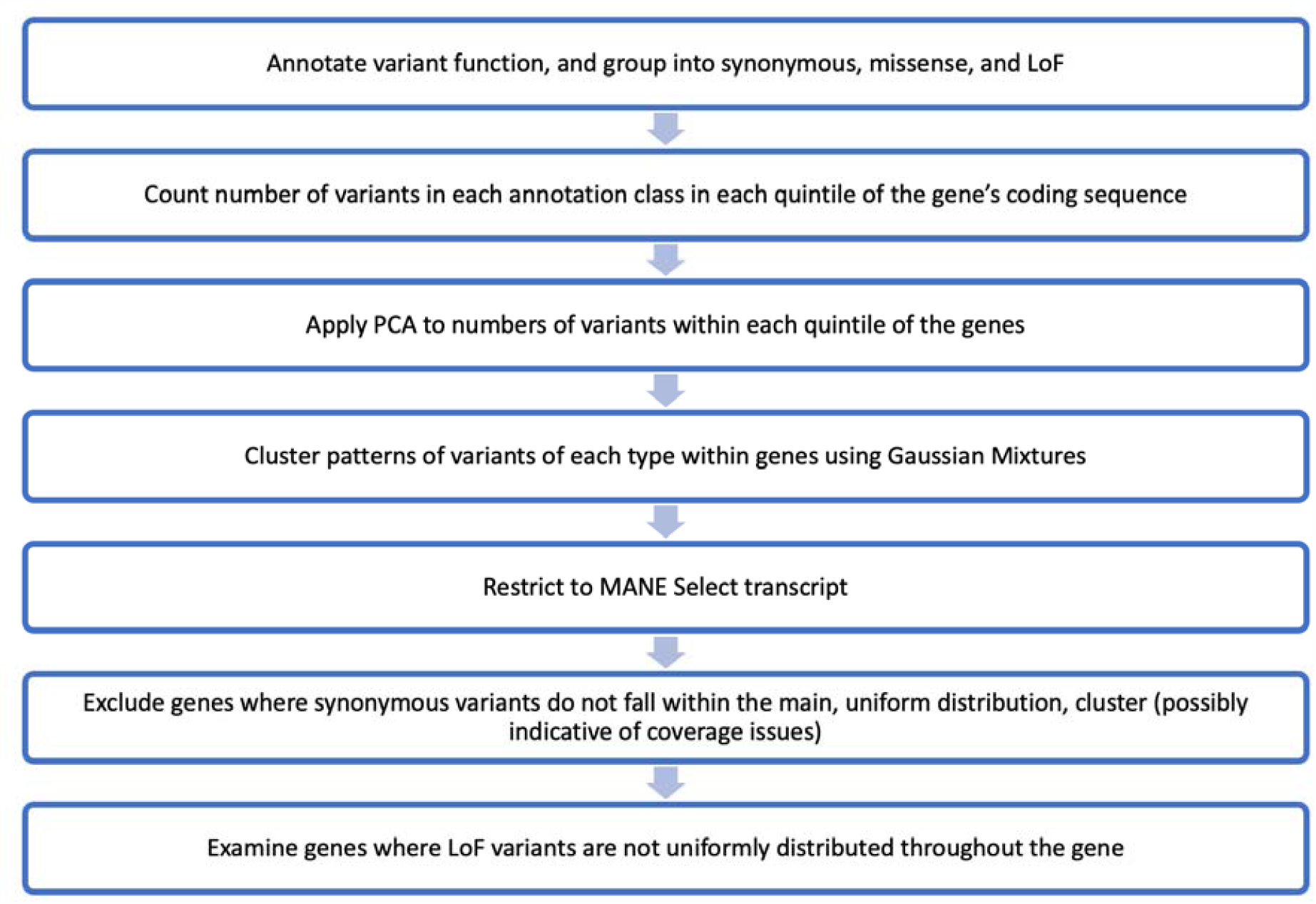
Flow diagram showing the experimental design.

**Figure 2:**
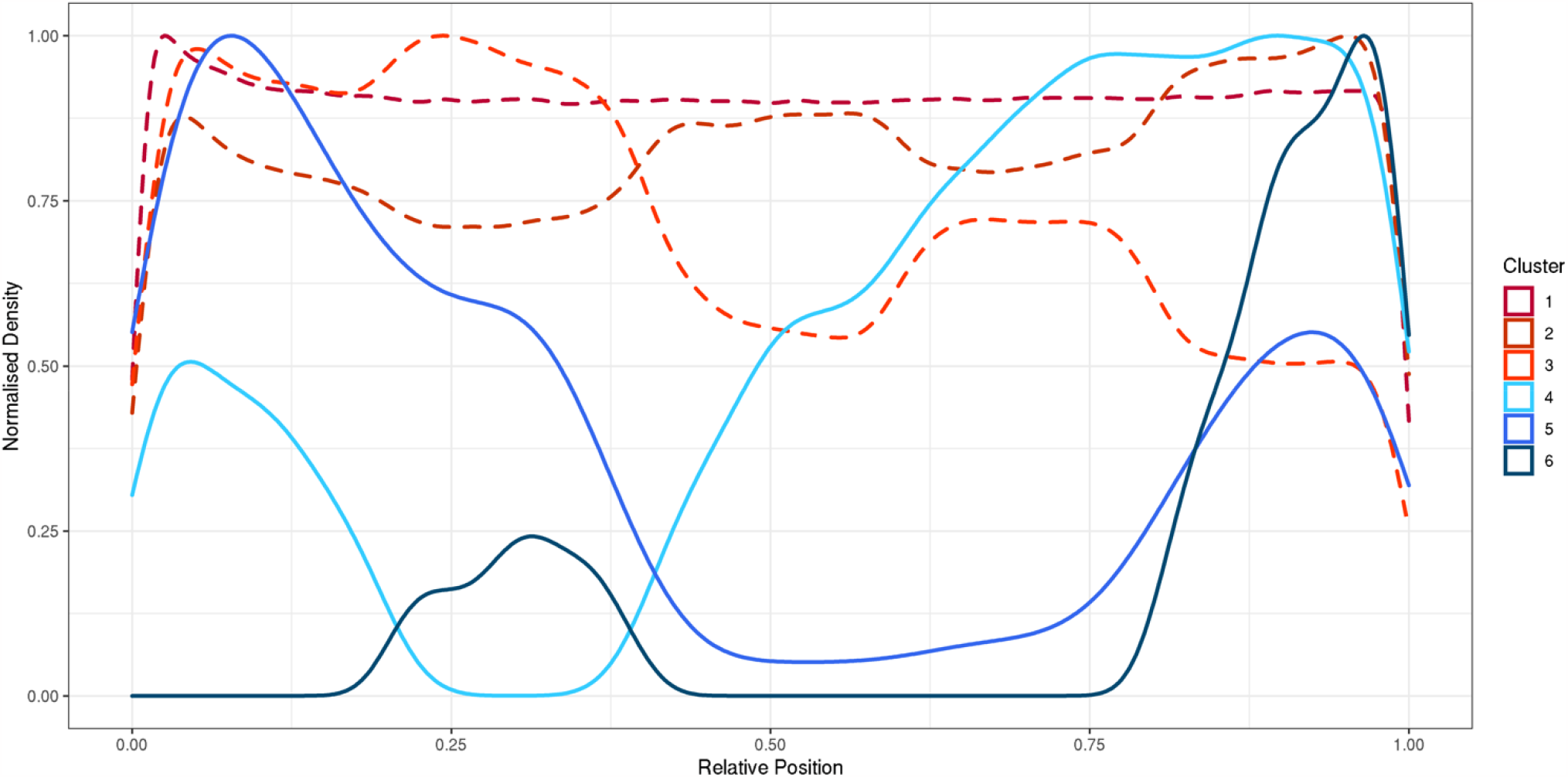
Profile of variant locations in each identified cluster. Frequency density plots of the relative position of all variants in all genes falling into each cluster. Shown are the 6 clusters which identified genes where variants of a particular class are present. The 7^th^ cluster identified genes where there were no variants of a given class.

We limited our analyses to transcripts with at least five variants of each consequence class, and only considered MANE Select transcripts for each gene in our primary analysis (16,473 genes). We found that, for most genes, synonymous and missense variants fell within the uniform clusters 1-3 (99.3% and 99.4% respectively; Fig. 3) and we excluded 114 genes with synonymous or missense variants in non-uniform clusters 4-6 from further analysis as these could be indicative of poor coverage over large regions of the gene. In contrast, we found considerably more genes with pLoFs in the non-uniform clusters 4-6 (n=1460, 8.9%) compared to synonymous and missense (P < 2.2e-16). These distinct profiles for pLoF location were not driven by the possible locations of pLoF variants, based on the underlying sequence, where only 63 genes had non-uniform distributions. Simulations showed that, while constrained genes were more likely to fall into the non-uniform distribution clusters, there was an enrichment of genes within these clusters compared to the expected distribution (p<0.0001; Fig. 4).

**Figure 3:**
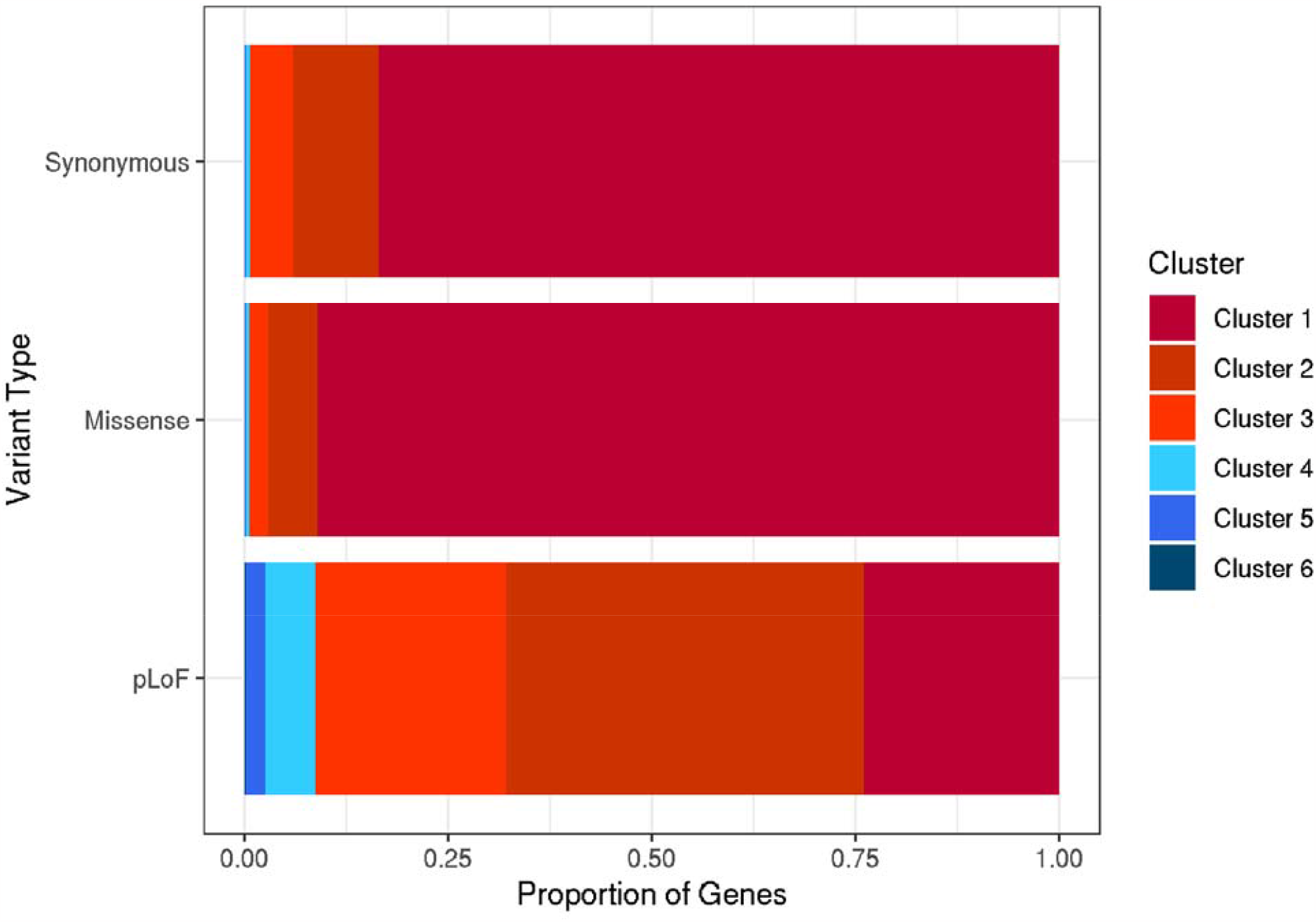
Proportion of genes falling into each cluster, separated by variant class. For each class of variant (synonymous, missense, pLoF) we present the relative proportion of genes where variants of that class were included in each of the 6 clusters where variants are present. The seventh cluster identifying genes with no variants of a particular class was excluded from the relative proportion calculations.

**Figure 4:**
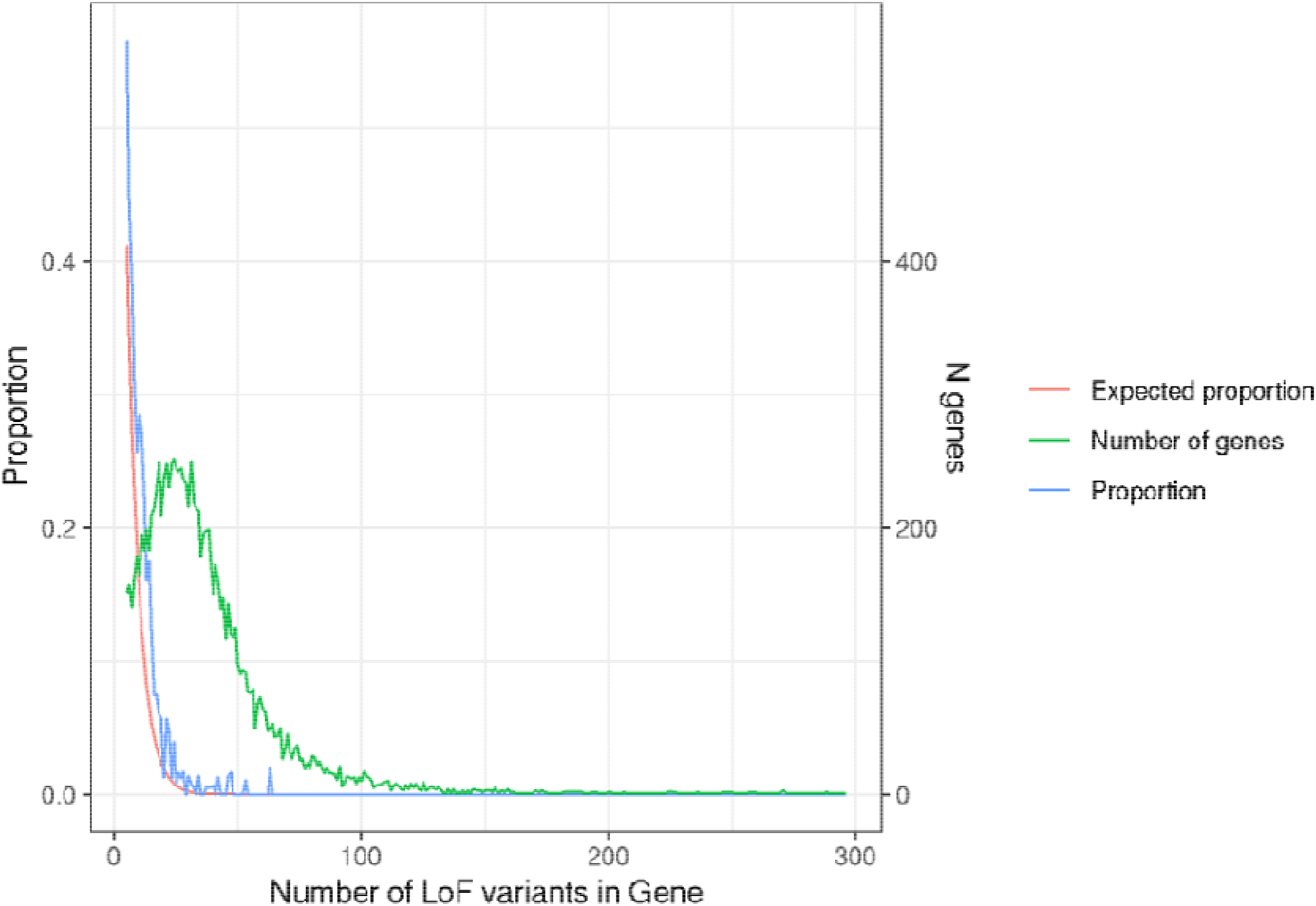
Proportions of genes in cluster 4-6 in UKB compared with simulations. The proportion of genes where pLoFs in UK Biobank are clustered into clusters 4-6 against the number of pLoFs in that gene is shown in blue. The proportion of simulated genes where pLoFs clustered into clusters 4-6 against the number of variants in the gene is shown in red. The green line shows the number of genes with each number of pLoFs in UK Biobank.

### pLoF variants are more likely to be non-uniformly distributed in genes linked with autosomal dominant conditions

We observed that pLoF variants in UKB were more likely to be non-uniformly distributed in autosomal dominant DD genes (AD-DD)^19^ versus other genes, possibly indicating regions where pLoFs are tolerated and do not cause severe disease. Of AD-DD genes, where pLoFs cause DD through haploinsufficiency (421 genes)^19^, we observed 29 with no pLoFs in UKB and 153 with <5 pLoFs (Table S1). Of the remaining 239 AD-DD genes, pLoFs in 41.4% were non-uniformly distributed (Fig. 5), which contrasted with 3.3% for autosomal recessive DD (AR-DD) genes. Genes linked with a range of adult-onset autosomal dominant diseases (including cardiac conditions, heritable cancer syndromes and eye disorders), were also more likely to have non-uniformly distributed pLoFs (25.3%) than other genes. Simulations showed that while these genes generally had fewer pLoFs than other genes, this did not explain the non-uniform distributions of pLoFs (p<0.0001).

**Figure 5:**
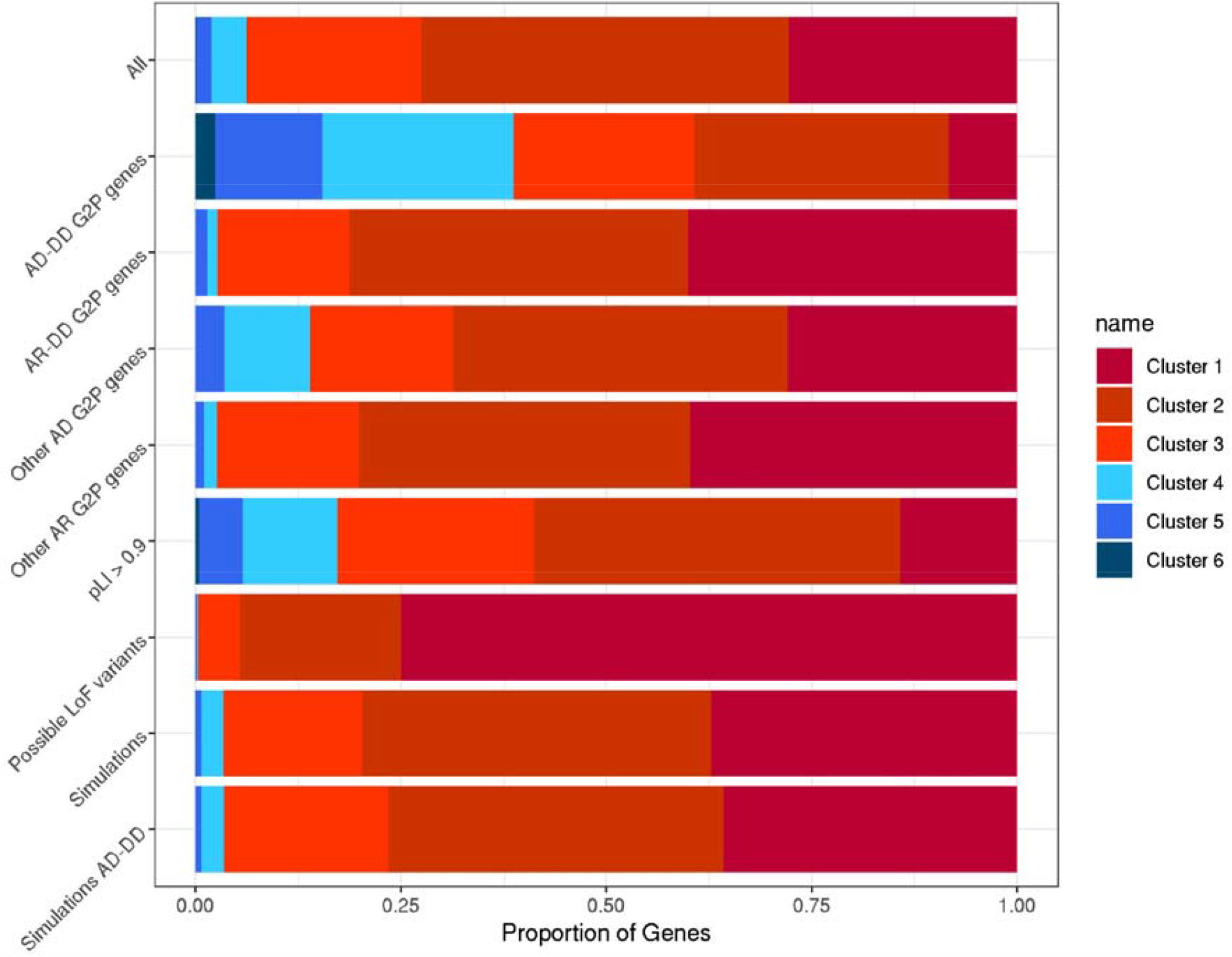
Proportion of genes with pLoFs in each cluster, including subsets of disease genes, locations of possible pLoFs, and simulation analyses. The relative proportion of genes where pLoFs are included in each of the six clusters with at least five pLoF variants is shown for different sets of genes, locations of possible pLoFs, and simulations: All (all genes with at least five pLoFs); AD-DD G2P genes (genes where pLoFs cause developmental delay through haploinsufficiency); AR-DD G2P genes (genes where pLoFs cause developmental delay through recessive mechanisms); other AD G2P genes (genes where pLoFs cause adult onset diseases, including cancer syndromes and heritable cardiac, eye or skin conditions); other AR G2P genes (genes where pLoFs cause adult onset diseases through recessive mechanisms); genes with high probability of LoF intolerance (pLI^15^) scores >0.9; possible LoF variants based on the underlying sequence of each gene; simulations of all genes (simulated genes matched to the number of pLoFs in each gene in UKB); simulations of AD-DD genes (simulated genes matched to the number of pLoFs in AD-DD genes in UKB). AD = autosomal dominant; AR = autosomal recessive.

Applying the same clustering procedure to disease causing (pathogenic/likely pathogenic) pLoF variants in ClinVar^20^ we found that among 1438 genes with at least five such variants, only 51 (3.5%) were uniformly distributed. Across 156 AD-DD genes with at least five pLoFs in both UKB and ClinVar datasets, we found 83 (53.2%) genes where pLoFs fell into one of the uniform clusters in UKB, compared with 150 (96.2%) in ClinVar (2-sided binomial P < 2.2x10^−16^). The majority of genes clustered similarly in both datasets; for example, pLoF variants in *COL4A3* (associated with Alport syndrome, MIM #104200) are uniformly distributed throughout the gene in both UKB and ClinVar (Fig. 6a). In such cases, where pLoFs are uniformly distributed throughout a gene in both population and clinical datasets, this approach is not able to determine why some pLoF variants are likely to be benign whilst others are pathogenic.

**Figure 6:**
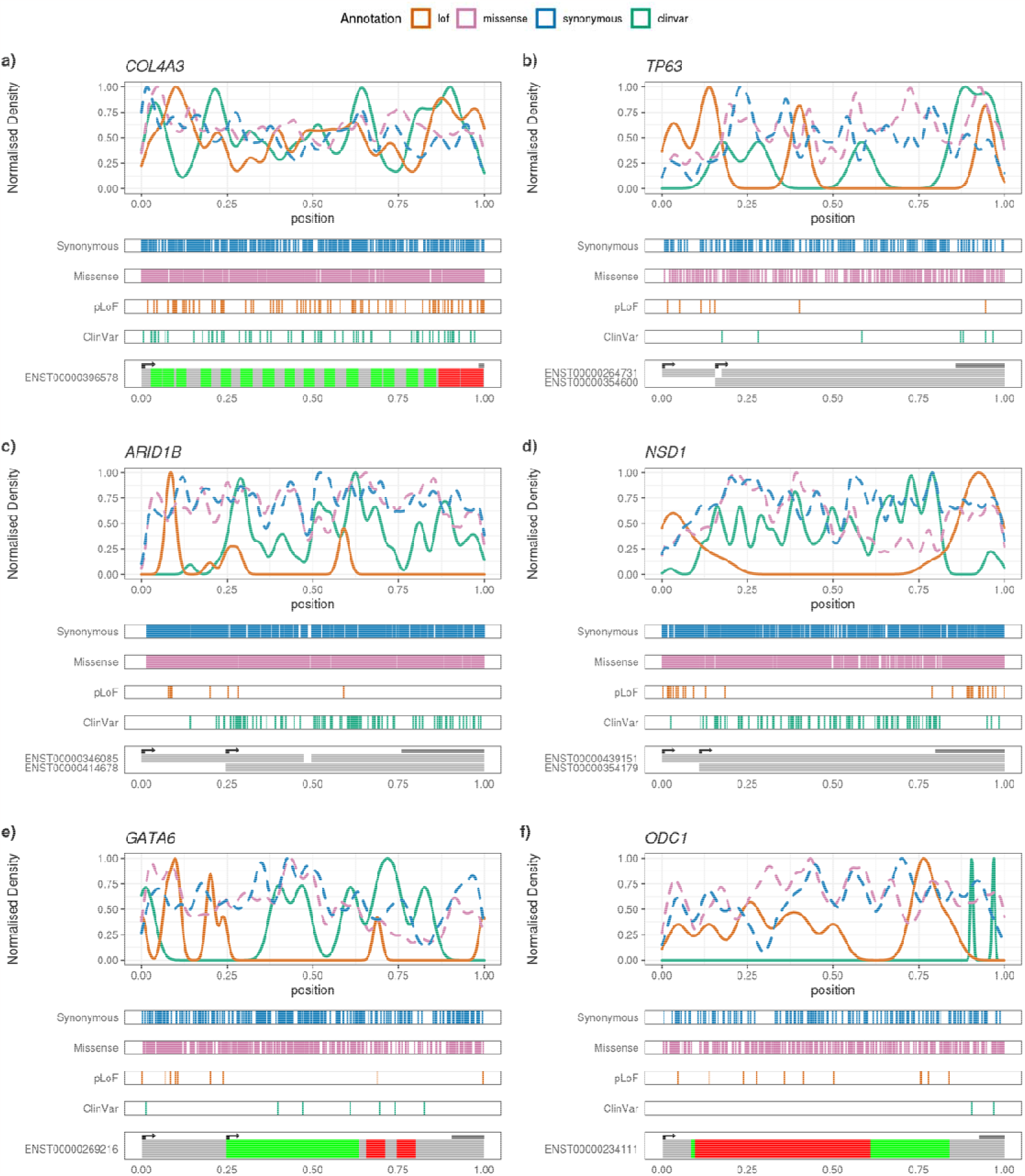
Profiles of variants of each class within selected genes Locations of variants of each class in UK Biobank individuals, and ClinVar pathogenic/likely pathogenic variants in *COL4A3, TP63, ARID1B, NSD1, GATA6*, and *ODC1* are shown. The top panel of each figure shows a frequency density plot of the relative position of variants of each class in UKB, plus ClinVar pathogenic/likely pathogenic variants. The middle panels show rug plots of the relative positions of each variant of each class in separate panels. The bottom panels show the locations of start codons, and a diagram of either the relative positions of domains within the protein (for *COL4A3, GATA6, ODC1*), or a depiction of the exons included in the labelled transcript (*TP63, ARID1B, NSD1*). The location of the final exon is indicated by the dark bar above the transcript diagram.

### Non-uniform distributions of pLoF variants may explain incomplete penetrance through a variety of molecular mechanisms including alternative splicing and translation re-initiation

In 43.6% of genes with at least five pLoF variants in both datasets, the distribution of pLoF variants differed substantively between UKB and ClinVar (i.e., one was uniform whilst the other was non-uniform). We hypothesised that these might represent examples where variant location could explain incomplete penetrance. We examined this list of genes for examples where the difference in distributions was robust, based on visual inspection of the underlying pLoF variant distributions and sequence data, and investigated potential mechanistic explanations.

One mechanism which might explain incomplete penetrance is the existence of alternative transcripts, where benign pLoF variants are clustered in exons that are excluded from other functional transcripts. For example, *TP63* (associated with Hay-Wells syndrome, MIM #106260) has seven pLoFs in the MANE Select transcript in UKB, of which five are in early exons not present in the MANE Plus Clinical transcript (Fig. 6b). This transcript (ENST00000354600) has an alternative later start codon but contains all the ClinVar pathogenic pLoF variants. In addition to MANE Plus Clinical transcripts, which may be the most obvious candidates for alternative transcripts to explain the presence of pLoFs in apparently healthy individuals, other transcripts may show higher expression levels and explain non-penetrance of some disease genes. Examples highlighted by our analysis include two large genes, *ARID1B* (associated with Coffin-Siris syndrome, MIM #135900) and *NSD1* (associated with Sotos syndrome, MIM #117550), which have 13 and 31 pLoFs in UKB respectively. For *ARID1B*, 10/13 pLoFs fall before Met584 of the MANE Select transcript (ENST00000346085), which also corresponds to the start of an alternative transcript (ENST00000414678) that shows higher expression in GTeX v7 than the MANE Select transcript^21^ (Fig. 6c). For *NSD1*, all the pLoFs in UKB occur either in the large last exon or the first exon of the MANE Select transcript (ENST00000439151) (Fig. 6d). The first exon is excluded from an alternative transcript, ENST00000354179, which has much higher expression levels in GTeX v7 than the MANE Select transcript. While the final exon is included in both transcripts, it lies downstream of the functional domains of the protein, and since pLoFs in the final exon usually escape NMD, a functional C-terminally truncated protein could be produced. In both cases, the pLoF variants in ClinVar are fairly uniformly distributed throughout the rest of the gene, but lie outside of these exons.

It is also important to consider not only alternative transcription but also translation re-initiation in explaining incomplete penetrance, as alternative start sites on the same transcript could rescue some pLoF variants (though these may not be annotated as such). For example, *GATA6* (associated with pancreatic agenesis and congenital heart defects, MIM #600001) has 10 pLoFs in UKB, of which eight are located before Met147 (Fig. 6e). This contrasts with ClinVar variants, which all lie after Met147.

Although there is only a single known transcript for this gene, *GATA6* can be produced through translation re-initiation from Met147, creating a second recognised protein isoform^22^ that is shorter but still retains the functional domains. Unlike the many other AD-DD genes, the phenotypes linked with *GATA6* haploinsufficiency are both specific and severe enough that we considered it implausible they would not be recorded in the linked electronic health records of UKB participants; importantly, we note that none of the 30 carriers have any indication of either pancreatic agenesis or cardiac malformations.

### Pathogenic pLoF variants at the end of genes may point towards a gain-of-function disease mechanism

Finally, we also found a small number of AD-DD genes where pLoF variants were uniformly distributed in UKB but non-uniformly distributed in ClinVar. For example, in *ODC1* (associated with Bachmann-Bupp syndrome, MIM #165640), all 11 pLoFs in UKB occur before the penultimate exon, whilst ClinVar pathogenic variants all occur in the last or penultimate exons (Fig. 6f). Here, despite being annotated as pLoF, there is no evidence that haploinsufficiency causes disease, and pathogenic variants at the end of the gene are likely to result in a gain-of-function (GoF), for example, by causing resistance to normal degradation^23^.

## Discussion

Using cluster analysis, we have identified 1460 genes which show distinct patterns of pLoF location within UKB, of which 16.4% are in genes where haploinsufficiency causes monogenic diseases that are generally assumed to be fully penetrant. We have also highlighted specific examples of well clinically characterised genes, including *GATA6* and *ARID1B*, where we were able to suggest potential molecular mechanisms that may explain the presence of pLoF variants in apparently healthy individuals. These examples show the importance of examining alternative transcription and alternative translation to understand the clinical impact of pLoFs.

Haploinsufficient genes can be divided into three groups based on the distribution of population genetic variation in UKB: (1) those where we observe too few pLoF variants to be able to cluster them effectively (37.4%); (2) those where we observe distinct non-uniform patterns of pLoF variant distribution (20.9%); and (3) those where we observe a broadly uniform distribution of pLoF variants (41.6%). For the first of these groups, the low numbers of pLoFs in UKB may be the result of haploinsufficiency in these genes being genuinely highly penetrant. For the second group we have demonstrated how this distribution can explain incomplete penetrance of pLoF variants in many of these genes. The final group of genes (where we observe uniform distributions of pLoFs in UKB) is perhaps most puzzling; although a subset may exhibit patterns of pLoF variant locations that are below the resolution captured by the quintiles used in our clustering approach, this is unlikely to be the case for all of them. Similarly, although a subset may cause unrecognised developmental disorders in some individuals, this is unlikely to be true for the majority given the known ascertainment bias towards healthy individuals in UKB^24^. However, some genes (such as *SRCAP*^25^) contain pLoFs that cause clinically distinct DDs via different mechanisms based on their location, with phenotypes ranging from mild to severe. For other genes with uniformly distributed pLoFs, the presence of incompletely penetrant pLoF variants may instead indicate the presence of modifiers, potentially in other genes or nearby non-coding regions. Understanding the mechanisms which modify the penetrance of these genes will require sequence data on large numbers of affected individuals to compare to healthy controls and is beyond the scope of this study, but would enable assessment of genotype-phenotype correlations and disease mechanisms at a sub-genic level.

While our study has identified genes where distinct patterns of pLoFs point toward mechanisms that may explain incomplete penetrance, there are some notable limitations. The use of quintiles to normalise the position of variants within genes means there will be patterns which are missed by our clustering approach, as their distribution is below the resolution captured by quintiles. Also, as demonstrated by the simulation analyses, there are a number of genes which will fall into clusters with distinct patterns of pLoF distribution by chance, rather than being driven by underlying biological mechanisms. Identifying these genes and separating them from those where the patterns of pLoFs are informative requires additional data and may not always be possible with high confidence. However, we believe that we have demonstrated the utility of our approach, which will improve with larger datasets. Additionally, while none of the individuals in UKB carrying pLoFs in the genes highlighted have been diagnosed with any of the conditions in question, there may be relevant phenotypes not captured in the UKB data. Increasing the sample size would also allow us to increase the robustness of the clustering, especially for highly constrained genes with few carriers in UKB.

We have shown how genes associated with assumed fully penetrant childhood-onset conditions through haploinsuffiency can have regions where predicted pathogenic variants are tolerated and don’t cause disease. Excluding such variants from both diagnostic pipelines and studies of disease penetrance is crucial. For example, within *GATA6* and *ARID1B*, we suggest that pLoFs occurring in the first quarter of the CDS of the MANE Select transcript (corresponding to the first 146 and 583 amino acids of the proteins, respectively) do not cause disease and should not be reported diagnostically. We have also demonstrated the benefits of using regional rather than gene-wide constraint metrics to understand the potential impact of pLoF variants, and our results may be helpful in determining whether pLoF variants in genes associated with monogenic conditions cause disease.

## Data Availability

All data produced in the present work are contained in the manuscript

## Acknowledgements

We thank Andrew Wood for assistance with the UK Biobank exome sequencing data, and members of the Exeter rare variant group for helpful feedback on this work. This research has been conducted using the UK Biobank Resource under Application Number 49847 and 9072. The authors would like to acknowledge the use of the University of Exeter High-Performance Computing facility in carrying out this work. This study was supported by the National Institute for Health and Care Research Exeter Biomedical Research Centre. The views expressed are those of the authors and not necessarily those of the NIHR or the Department of Health and Social Care. This work was supported by the Medical Research Council [MR/T00200X/1].

## Methods

### Classification of variants in UK Biobank

Variants from exome sequencing (ES) data within the UK Biobank were called centrally by the UKB team using graphTyper^26^. We used the Ensembl VEP v104^18^ with the LOFTEE plugin^14^ to annotate the variants with their predicted functional consequences. We excluded variants which were flagged for removal by UKB due to low depth. Within each protein-coding transcript, we grouped variants into synonymous (VEP classification “synonymous_variant”), missense (“missense_variant”), and pLoF (variants classified by LOFTEE as “LoF”). No frequency threshold was used, and variants were all counted once regardless of minor allele frequency. We further separated pLoF variants into SNVs, and frameshift insertions or deletions (indels). For our main analyses we considered all pLoF variants together; pLoF SNVs were used as a sensitivity analysis to ensure patterns were not driven by indels spanning quintiles of the gene. Using start and end locations for exons within each transcript from Ensembl (downloaded on 01-04-2022), we calculated the relative position of each variant within the coding sequence of each transcript, considering only exonic variants (i.e., excluding splice donor and splice acceptor variants). We then divided each transcript into quintiles, and for each class of variants (synonymous, missense, pLoF) we calculated the number of variants within each quintile as a proportion of the number of variants of that class.

### Cluster analysis

We applied principal components analysis (PCA) to the proportion of variants in each quintile of the gene transcript, separated by variant class. PCA loadings were calculated based on locations of synonymous, missense, and all pLoF variants. We then projected the PCA onto SNV pLoFs, possible pLoFs, simulations, and ClinVar variants.

We clustered the PCA profile of variants of each class within each transcript using Gaussian Mixtures, allowing seven clusters, trained using the profile of synonymous, missense and all pLoFs. For each class of variant within each transcript we obtained the most likely cluster, as well as the probability for its inclusion in each cluster. We projected the SNV pLoFs, possible pLoFs, simulations and ClinVar variants into the clusters to obtain their most likely cluster and probabilities. Seven clusters were chosen to allow for multiple clusters with different non-uniformly distributed variants. We performed sensitivity analyses varying the number of clusters to ensure that our results were robust to the number of clusters chosen.

### Sensitivity analyses

#### Possible locations of pLoF variants

We examined the coding sequence of each transcript and calculated the locations of all possible pLoF SNVs. We clustered these in the same way as observed pLoF variants (see below) to verify that any patterns we identified were not driven by the underlying coding sequence, and the possible locations of pLoF SNVs. The clustering of these variants was compared to that of observed pLoF SNVs.

### Simulations

To estimate the rate at which genes with a given number of variants of a given class clustered into each cluster, we used simulations to create synthetic sets of genes with varying numbers of variants. We took the relative positions of all variants in the UKB ES data, and randomly selected a number of these based on the number of pLoF variants within each gene in UKB. We repeated this 10,000 times. Simulated genes were then clustered to estimate the number of genes falling into each cluster by chance.

### ClinVar variants

We downloaded clinically annotated variants from ClinVar (02/10/2022) and calculated the proportion of pathogenic/likely pathogenic pLoF variants within each quintile of each transcript and clustered them to compare to the UK Biobank pLoF clusters.

### Disease gene lists

We examined the clusters which pLoFs fell into in genes linked with monogenic diseases from the Gene2Phenotype database to investigate whether these could elucidate the variable penetrance of these genes. Gene lists were downloaded from https://www.ebi.ac.uk/gene2phenotype/ (accessed 04-06-2021) and split into those causing severe developmental disorders (DD), and those causing later onset diseases (cancer, cardiac, eye, and skin). These were further subdivided into monoallelic (autosomal dominant) and biallelic (autosomal recessive) genes with “absent gene product” mechanisms; G2P genes with other inheritance classes or disease mechanisms were excluded.

